# A collaborative regional multisite and multi-departmental initiative for improving consensus on clinical outcomes data in spine surgery

**DOI:** 10.1101/2025.06.09.25329269

**Authors:** Bahar Shahidi, Robert K. Eastlack, Gregory M Mundis, Choll Kim, R. Todd Allen, Steven R. Garfin, Joseph Osorio, William Taylor, Peter O. Newton, Samuel R. Ward, Behrooz A. Akbarnia

**Author notes:** CORRESPONDING AUTHOR: Bahar Shahidi, PT, PhD, 9500 Gilman Dr. MC0863, La Jolla CA 92093.

## Abstract

**Study Design:** Retrospective chart review with consensus.

**Objective:** To establish a consensus-driven multisite collaborative database of individuals undergoing care for spondylolisthesis as a model for identifying barriers to standardized quality data collection in spine care.

**Summary of Background Data:** As health care initiatives move toward value-based care implementation, improving standardization of data collection through large databases and registries is increasingly important. However, in the absence of resources, many community practices are not represented, and models for low-cost implementation of such data collection efforts have yet to be established.

**Methods:** Data sharing agreements were established across participating private, community, and academic spine centers and data from charts of patients with spondylolisthesis was retrospectively reviewed for prevalence of demographic, surgical, operational, and outcomes variables. A consensus-based needs assessment then identified sources of error, additional data needs, and barriers to collection.

**Results:** 115 adult and pediatric charts were reviewed across 10 surgeons and 4 sites. High prevalence (>90%) variables included demographic (age, gender, BMI, smoking status) and operational variables (operating room time, blood loss, length of stay, 90-day readmission rate), and preoperative back pain severity. Low prevalence variables (<25%) included medication use, prior treatments, and skin to skin time for staged surgeries. 90% of surgeons identified inconsistencies between the coded procedure documented and surgery type. Variables identified for future collection included leg pain severity, and a single measure for comorbidity assessment. Barriers to quality data collection included lack of administrative resources, education of research or medical assistants extracting data, and regulatory burden.

**Conclusion:** These data provide a model for a low-cost collaborative database infrastructure, with recommendations for standardized data collection. Errors and site variability in surgical coding may have a significant impact on interpretability of study designs involving chart review, along with sparsity of patient reported outcomes.

## INTRODUCTION

Over the past decade, there has been an effort to improve standardization of health care data collection to optimize patient care, follow up, documentation, and care reimbursement(1). Uniformity could help facilitate collaboration between community and academic settings on projects aimed at improving patient care quality, safety, and value. Further generating centralized and accurate data through which treatment outcomes can be compared is a powerful tool for studies on treatment efficacy and effectiveness. Creating a model system for standardized outcomes in a population- and clinic-specific manner for clinicians specializing in treatment of spine conditions provides the foundation for widespread implementation.

Degenerative and isthmic spondylolisthesis are among the most common pathologies for which patients seek care in the spine(2). Despite this commonality, nonoperative treatment and surgical techniques utilized to manage the pathoanatomy of spondylolisthesis vary widely amongst specialists(3). Such disparity in treatment potentially results in considerable variance in costs, complications, and clinical outcomes. The effect of surgical and nonsurgical treatment differences on these variables remains poorly understood. Additionally, the burden on community clinical practices of collecting and organizing outcome measures is often too high without informed structural guidance and resources from larger academic research partners. To develop data collection infrastructure and opportunities for representation of clinical data across both community and academic settings involved in treating spine conditions such as spondylolisthesis, an evaluation of the operational barriers and data availability is necessary. To accomplish this, we applied a consensus-based approach to data collection and quality assessment representing a spectrum of spine practice settings for both adult and pediatric practitioners treating spondylolisthesis. The purposes of this project were to: 1) characterize current practices for clinical data collection across various spine practice settings and practitioners, 2) identify barriers to implementation of a multisite collaborative database for these settings, and 3) evaluate future needs for data collection with the goal of standardizing collection guidelines across spine care practitioners.

## METHODS

### Shareholder recruitment and database infrastructure

Surgeons from local spine centers were invited to social and academic meetings to encourage participation and commitment to the process. There was an effort to represent a diverse spectrum of providers, including private practice, community-based institutions, and academic institutions/teaching hospitals that reflects observations of current national practice patterns(4). Similarly, both neurosurgical and orthopaedic surgery spine specialists across various patient populations were invited to participate. In the initial stage, interested participants met monthly to establish a comprehensive list of variables commonly collected as part of standard clinical and surgical practice for the patient populations treated. The chosen variables were inclusive of those achievably captured by all sites, and of potential merit in identifying differences in various utilization or outcome metrics. Once the desired list of variables was finalized, individual ethical review board approvals were obtained on a per-site basis for chart review and data collection. Data sharing agreements across sites and a database infrastructure were established using a standardized case report form within REDCap, housed at a single academic institution. The case report form included variables that could potentially be collected across all sites, to evaluate the quality and quantity of data across treatment settings.

### Data Collection and Consensus Review

Participating surgeons were surveyed regarding patient populations treated, length of follow up, and medical record system utilized. Charts from 20-30 patients per site with single level degenerative or isthmic lumbar spondylolisthesis were retrospectively reviewed for prevalence of demographic, surgical, operational, and outcomes variables. Based on the prevalence of data for each variable collected, a data review was performed on a per/surgeon basis to summarize results, check for errors in data, and identify missing data. Prevalence of data errors, missing variables, and inconsistencies between data collected and the surgeon’s interpretation of the treatment were documented. A consensus-based needs assessment was then performed via group discussion and surveys to share results of the data collection and perform data quality checks. Finally, recommendations were made based on majority agreement for priority focus areas for improvements in data integrity, and for variables of clinical interest that should be added to standard data collection protocols (Figure 1).

**Figure 1.**
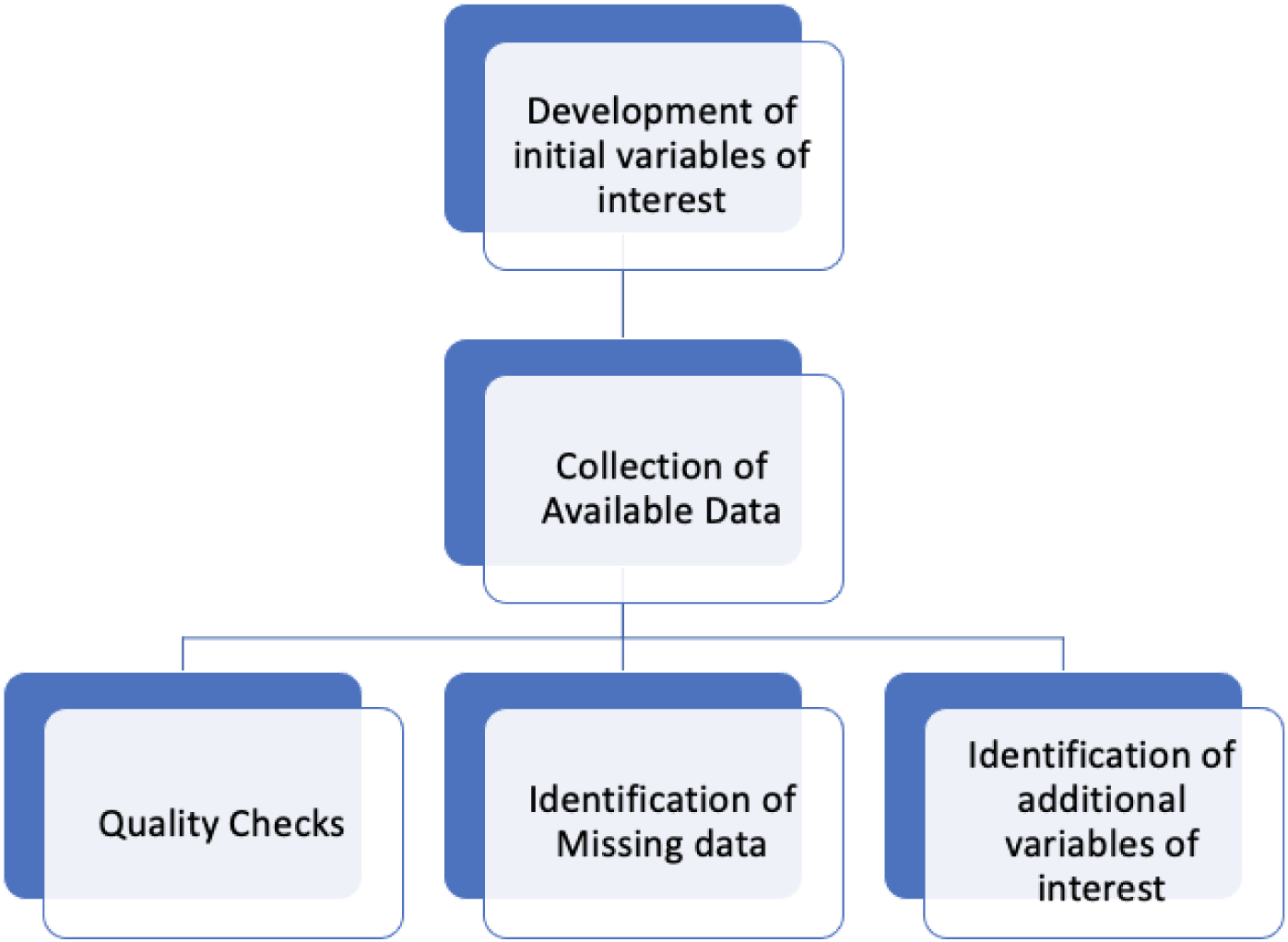
Conceptual framework for implementation of consensus-based multisite data collection infrastructure.

## RESULTS

### Shareholder Recruitment

Of 15 initial invited providers, 10 surgeons agreed to participate in data collection. Of surgeons who opted out of participation, 4 (80%) did not participate due to lack of administrative support for data collection (from private practice settings), and 1 (20%) did not participate due to burden of establishing regulatory permissions (from community-based setting). The 10 participating surgeons represented 4 different sites, including 1 private practice, 2 community-based institutions, and 1 academic institution. 80% of participants were orthopaedic surgeons, and 20% were neurosurgeons. 90% of participating surgeons treated primarily adult populations, and 10% treated primarily pediatric populations.

### Data Collected

Sequential charts for 95 adult and 20 pediatric patients were reviewed. Variables included for initial collection after the first consensus review are listed in Table 1. Variables with >90% collection rates across sites included age, gender, body mass index (BMI), smoking status, diagnosis, surgery type, operating room (OR) time, estimated blood loss (EBL), discharge disposition, length of stay (LOS), 90-day readmission rate, and pre-operative back pain severity (VAS). Variables with <25% collection rates across sites included medication use, prior treatments, skin to skin time for each stage of a staged surgery, and post-operative disability (Oswestry Disability Index) scores at any timepoint. From the reported data, surgical treatment approaches for spondylolisthesis varied according to surgeon preference and patient population (Table 2).

**Table 1.** List of variables included in case report form for initial collection.

| Demographic | Surgical | Outcomes |
| --- | --- | --- |
| Age (years) | OR time | 90-day readmission |
| Sex (M:F) | Skin to Skin time | Length of Stay |
| Body Mass Index | Estimated Blood Loss | Discharge Disposition |
| Comorbidities | Procedure type | Post-operative VAS* |
| Occupation/Sport | Fixation type | Postoperative ODI* |
| Smoking status | Intraoperative Corticosteroids | Medication Use |
| Spondylolisthesis level | Neuromonitoring | * collected at 3, 6, and 12 months postoperatively |
| Spondylolisthesis type | Drain usage/duration |  |
| Medication usage | transfusions |  |
| DEXA |  |  |
| Prior treatment |  |  |
| Baseline VAS |  |  |
| Baseline ODI |  |  |

**Table 2.** Patient characteristics for surgical procedures performed on the patient population. *32% of pediatric direct fixations were also coded as Open Reduction Internal Fixation (ORIF)

| Variable | Adult | Pediatric |
| --- | --- | --- |
| Age: Mean (SD) years | 63(11) | 15(2) |
| Spondy type (%Degen) | 91% | 0% (all isthmic) |
| Majority Spondy Level | L4-5 (61%) | L5-S1 (93%) |
| Surgery Type |  |  |
| ALIF | 18% | 0% |
| LLIF | 46% | 0% |
| TLIF | 25% | 3% |
| PLF/PLIF | 60% | 47%* |
| Decompression | 51% | 61% |
| OR time: Mean (SD) min | 238(280) | 282(100) |
| Length of Stay (days) | 2.5(3) | 3(1) |
| Preop ODI | 31(18) | NA |
| Preop VAS | 6.3(3) | 1.3(2) |

In the pediatric population, the most common surgical approach was laminectomy with posterior fixation (53.6%). In the adult population, surgery type varied by site, with lateral approach fusions being most common (18-75%). However, this data contained errors due to heterogeneity in coding and interpretation, with 90% of surgeons identifying inconsistencies between the coded procedure and type of surgery performed. Prevalence of patient reported outcomes (PROMs) for back pain and disability varied by site, duration of follow up, and patient population (adult vs pediatric). Back pain severity was collected in 70% of cases at 3 months, but only in 45% by 1 year. Disability was collected in 16% of cases at 3 months and remained consistent over 1 year. It was not collected in pediatric patients. Medication use, prior treatments, and skin to skin time for staged surgeries was available for 3-23% of cases (Figure 2).

**Figure 2.**
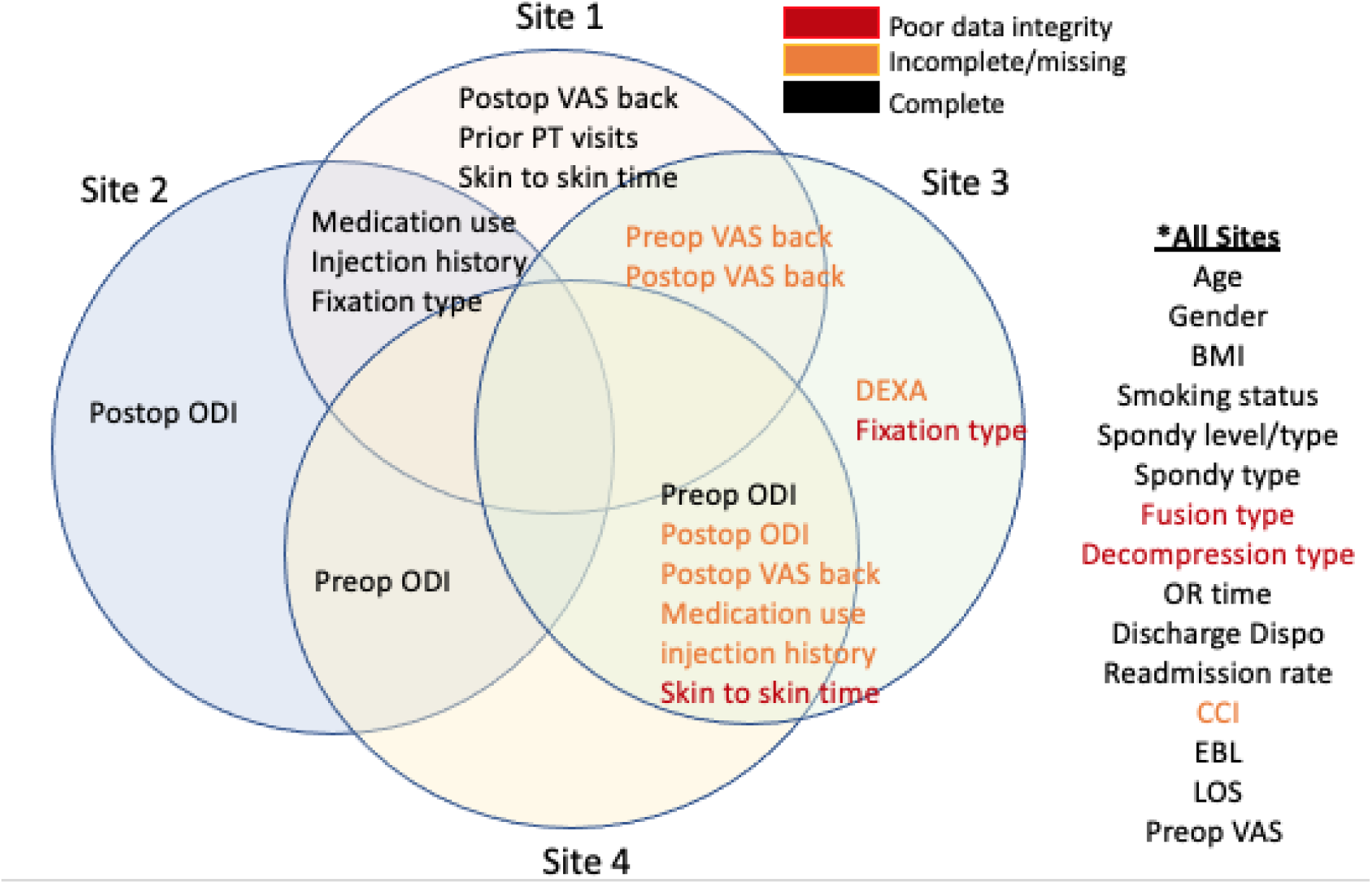
Results for data collected across sites. Variables with poor data integrity are indicated in red, variables with low prevalence are indicated in orange, and data that were collected with high prevalence are indicated in black.

### Consensus for Improvement and additional variables

After initial data extraction and review, consensus was reached (all surgeons were in agreement) for areas in which data integrity and quality was inadequate, and a recommendation was made on additional variables holding clinical value to collect. Of variables with recommendations for improvement, coding and surgical type definition, and documentation of skin-to-skin time for staged surgeries were identified as high priority items. Specifically, it was noted that there were some discrepancies in procedural codes across institutions for a similar surgery. It was also noted that for sites that utilized non-surgeons for data entry (e.g. research assistants, medical assistants), interpretation of surgical details was inconsistent and resulted in data entry errors. In regards to variables identified as valuable additions to standard clinical assessment and documentation, leg pain severity (for adult patients only) and a single metric for comorbidity assessment were recommended.

## DISCUSSION

Improving collaborative, centralized, and coordinated efforts to collect and analyze information related to patient healthcare in surgical outcomes has been highlighted as a critical need in many areas of medicine, including spine care(5). Although these efforts have been implemented on larger scales through databases and registries(5), feasible implementation often requires substantial resource investment, as well as collaborations with public health agencies, federal, and regulatory services. The goal of this project was to evaluate feasibility, consensus, and quality of a small-scale local collaborative registry and to identify key barriers to implementation across a spectrum of community providers, patient populations, and institutional infrastructures. We found that, with buy-in of participating centers and stakeholders, such an effort is possible with minimal resource utilization, although key barriers included lack of administrative help, regulatory burden, inconsistencies with data integrity, and variability in data collected across sites. Identification of these barriers highlights a need for infrastructural support to encourage better representation of accurate data from diverse provider groups and patient populations in larger-scale registries used for establishment of evidence-based practice guidelines.

Current national and international databases are becoming more common to meet the growing need for complete data on health practices for spine-related procedures. These databases have historically involved large institutions with substantial administrative and technological infrastructure, collaboration with public health centers of excellence, and commitment of resources associated with these services. Successful examples of such databases include the American Spine Registry (ASR)(5), the Quality Outcomes Database (QOD)(6), the International Spine Study Group (ISSG), and the Harms Study Group (HSG). Importantly, these national and international databases provide key data from which a large body of literature has evolved. As a result, they make a substantial impact on informing evidence-based practice recommendations and care implementation for individuals with spine conditions.

One observation from this smaller-scale collaborative effort was that there are inconsistencies in data sharing and regulatory considerations regarding human subjects’ research across institutions and regions. Ethical and regulatory requirements and safekeeping of patient health information (PHI) continues to evolve as new challenges in data safety emerge. Regulatory infrastructures and permissions developed over a decade ago for some databases or registries may not reflect current landscapes, and lack of administrative expertise to keep up with such adaptations are a likely to be a barrier for database development in under-represented and under-resourced practice settings(7). Reference to use of standards for interpretation of the Common Rule and definitions of PHI under HIPAA as suggested by the HHS OHRP (http://www.hhs.gov/ocr/privacy/hipaa/understanding/summary/index.html) and further clarified by current registries is encouraged when such regulatory barriers are encountered (7, 8).

Another surprising finding was the lack of consistency in both surgical approach, and surgery specific information as it was entered into the database when compared to the surgeon’s interpretation of the procedure. Some reasons for discrepancy in data acquisition may include variability in resources available for administrative support across sites, either in the form of professional and trained coders familiar with spine practice, or in the form of trained research or medical assistants who interpret operating room notes and enter surgical details in the case report form. Appropriate training and expertise of the administrative help tasked to data interpretation and entry is essential for data integrity and reduction of errors. Inconsistencies in coding and surgical data across sites have long been described as an important challenge in the context of standardizing procedural, billing, and reimbursement practices(9, 10). Additionally, differences in reimbursement and salary incentives between specialties as well as between private and academic institutions has been shown to influence practice patterns, that may further contribute to variability in surgical data and outcomes(11). However, the implications of this variability in practice and data integrity for research and quality assessment are less frequently addressed. Further research is needed to impact of these factors on generating and recommending standard practice guidelines, however high priority should be put upon appropriate training and oversight of administrative staff involved in interpretation and entering of data extracted from medical records.

Finally, we observed that although all clinicians were in consensus that PROMs were a high priority to collect, the prevalence of their collection was often lower than acceptable for reasonable data quality assessment, or evaluation of clinical outcomes at a level of rigor required by current research standards. However, safety and operational data such as length of stay, readmission rate, and OR time that are considered high priority for hospital administration and cost optimization were consistently collected. As the pressure for value-based care continues to increase, it is likely that recognizing, developing, and supporting infrastructure for collection of PROM’s at the level of business operations will become increasingly essential for progression of the field. Establishing a standardized data collection “toolbox” to be administered at each patient encounter may be an effective method for improving data acquisition, although future research is needed to evaluate the most efficient and cost-effective methods of data collection in a manner that is accessible to all practice settings.

## CONCLUSION

A successful regional multisite collaborative data sharing infrastructure was established for individuals with single level lumbar spondylolisthesis. Despite high data collection rates, errors associated with lack of sufficient training of data entry personnel, as well as site variability in surgical coding may have a significant impact on interpretability of data involving retrospective chart review, gathered between multiple surgeons/institutions, including large multisite registries. Additionally, sparsity of PROMs highlights a need for improving standardized data collection procedures for long term follow up.

## Data Availability

All data produced in the present study are available upon reasonable request to the authors

## ACKNOWLEDGEMENTS

This project was funded in part by the Scripps Clinic Medical Group. All study procedures were performed in concordance with the Declaration of Helsinki and under approval of each institution’s respective ethical review board.

